# Genome-wide analysis of tandem repeat variation identifies *SLC15A4* as a susceptibility gene for idiopathic pulmonary fibrosis

**DOI:** 10.1101/2025.11.25.25340948

**Authors:** John W. Oketch, Asalah Albahimah, Amilcare Barca, Richard Allen, Nick Shrine, Bruno Walewski, Ozge Sidekli, Afra Alyassi, Alessandro Romano, R Gisli Jenkins, Toby M. Maher, Philip L. Molyneaux, Iain Stewart, Tiziano Verri, Louise V. Wain, Edward J. Hollox

## Abstract

Idiopathic pulmonary fibrosis (IPF) is a progressive and fatal interstitial lung disease with a strong genetic component, yet a substantial proportion of its genetic risk remains unexplained by single-nucleotide variant (SNV) association studies. In this study, we conduct a genome-wide association study of tandem repeats (TRs), which are a pervasive and understudied class of genetic variation, to identify novel loci influencing IPF susceptibility. Using whole-genome sequencing data from two case-control datasets (discovery: 507 cases and 174 controls; replication: 1243 cases and 3197 controls), we identify and replicate associations at four TR loci, most notably a complex repeat within intron 2 of *SLC15A4*, a gene encoding the endolysosomal peptide-histidine transporter 1 (PHT1). This TR is within a predicted enhancer and acts as an expression and splicing quantitative trait locus (eQTL/sQTL), influencing the expression of an alternative *SLC15A4* transcript in biologically-relevant tissues and cells, with longer TR alleles correlating with decreased expression of this alternative transcript and increased IPF risk. In a biologically-relevant cell-line model, we show that this alternative transcript is targeted by nonsense-mediated mRNA decay, and levels of the alternative transcript are inversely correlated with levels of the canonical full-length *SLC15A4* transcript.

Our findings implicate *SLC15A4* in IPF pathogenesis, possibly via modulation of innate immune responses to injury, and demonstrate the power of TR-focused genomic analyses to reveal previously undetectable disease mechanisms and therapeutic targets.

## Introduction

Idiopathic pulmonary fibrosis (IPF) is an interstitial pneumonia characterised by progressive scarring of the alveoli, with a poor prognosis. It is a rare disease, with an incidence ranging between 0.76 to 7 per 100,000, increasing with age ^1^. Both environmental and genetic variation contribute to the risk of IPF. Genome-wide association studies of IPF susceptibility have identified 32 loci with a range of effect sizes, with the airway mucin gene *MUC5B* showing the largest odds ratio of five ^2–10^. As well as host defence mucins such as *MUC5B* and *MUC2*, other genes implicated by large-scale GWAS include those involved in telomere maintenance (e.g. *TERT*), cell-cell adhesion (e.g. *DPP9*), signalling (e.g. *DEPTOR*), and mitotic spindle assembly (e.g. *MAD1L1*). This reflects our current understanding of the etiological basis of IPF, which is an irreversible fibrosis as an inappropriate response to lung injury caused by, for example, viral infection or exposure to particulates by smoking ^11^.

GWASs focus on associations with single nucleotide variants (SNVs), but other genetic variation remains poorly captured by a GWAS, prompting approaches to analyse these other forms genome-wide and at scale ^12^. Variation at DNA tandem repeats is extensive throughout the genome, and both short tandem repeats (STRs) with repeat units 1-6 bp, or variable number tandem repeats (VNTRs) defined as repeats 7-100 bp, are an extensive source of genetic variation ^13–16^. Tandem repeat (TR) variation remains underexplored in disease studies because of two main reasons. Firstly, TR variation is not detectable by SNV-chip approaches, so current genome-wide approaches rely on high-coverage short-read sequencing, together with specialized analysis methods. Secondly, TRs are often not in strong linkage disequilibrium with flanking SNVs, so current imputation approaches are not completely effective ^17^.

Although the contribution of TRs to most complex diseases remains unclear, there is good evidence that extended alleles of particular TRs contribute to the risk of complex neurodegenerative diseases such as amyotrophic lateral sclerosis and frontotemporal dementia ^18–20^. There is also evidence that TRs influence human phenotypes more broadly ^21,22^. Furthermore, the role of TRs in causing Mendelian genetic diseases such as Huntington’s Disease and Fragile X syndrome is well-established ^23^.

In this study, we assessed the contribution of TR variation to IPF susceptibility. We used a dual approach to assess TR variation by using both GangSTR, which genotypes TRs based on a known catalogue, and ExpansionHunter Denovo (EHdn) which focuses on rarer TR expansions that may not be in a catalogue of known TRs. We applied these approaches to independent discovery and replication datasets, investigated the effect of associated loci on gene expression levels, and for the strongest association with *SLC15A4,* used experimental evidence to show the relationship between the disease-associated TR, transcript usage, and the role of nonsense-mediated mRNA decay (NMD) in controlling gene expression. Taken together, this work suggests *SLC15A4* as a novel, potentially druggable, locus for IPF.

## Methods

### Discovery dataset samples

For cases, 541 PCR-free whole genome sequences (WGS) from PROFILE (the Prospective Study of Fibrosis in the Lung Endpoints, ClinicalTrials.gov identifiers NCT01134822 and NCT01110694) study in the UK were analysed ^24^. These samples had been paired-end sequenced using Illumina Novaseq at an average genome coverage of ∼30x ^25^. An Amazon Web Services (AWS) cloud compute platform running Illumina DRAGEN Bio-IT Platform Germline Pipeline v3.0.7 was used to align the reads to the GRCh38_full_analysis_set_plus_decoy_hla.fa reference genome. These samples had been previously included in GWAS of SNVs^4^.

For controls, 292 control samples were obtained: 249 PCR-free WGS from the Genotype Tissue Expression Project (GTEx, accession phs000424.v8.p2, study number 29173) from the National Institute of Health (https://GTExportal.org/home/) and 43 PCR-free WGS from the Personal Genome Project Canada (PGPC) ^26^ - (https://personalgenomes.ca/). For both PGPC and GTEx samples, raw unaligned sequence reads in fastq files were retrieved. Low quality reads (i.e., reads with adapter carry-overs and those with a phred score<30) were removed using Trimmomatic software ^27^. The trimmed reads were aligned to the GRCh38 human reference genome using Illumina DRAGEN software, as for the cases. All the code and software used in this study are provided in the code availability and software section, and the data quality control steps are summarized in Supplementary Figure 1.

To assist with filtering of samples of non-European ancestries from the case-control datasets using TR genotype calls, an additional 102 PCR-free WGS were obtained from the open access 1000 Genome Project (1KGP). The 1KGP samples were selected to represent global variation: African (AFR; n = 26), European (EUR; n = 25), East Asian (EAS; n = 16), South Asian (SAS; n = 15) and admixed American (AMR; n = 20).

### Replication dataset samples

To validate the discovery of TR loci associated with IPF susceptibility, independent case-control datasets comprising of 1,473 individuals with IPF and 4,448 unaffected individuals sourced from the Trans-Omics for Precision Medicine (TOPMed) were utilized ^28^ (dbGAP study 34964). IPF-affected individuals (n = 1,473) were enrolled in the Familial and Sporadic Idiopathic Pulmonary Fibrosis study (Table 1). Because the TOPMed dataset does not include healthy controls, individuals unaffected by IPF were selected from the Cardiovascular Health Study (CHS) which recruited individuals older than 65 years. Depending on the TOPMed recruitment phase, samples were Illumina paired-end sequenced at different centers to at least a minimum of 30x coverage using a PCR-free protocol. A detailed report on library preparation, sequencing and alignment is reported here https://TOPMed.nhlbi.nih.gov/TOPMed-whole-genome-sequencing-methods-freeze-9 by the TOPMed consortium. All raw sequences were mapped to the human reference genome GRCh38 using BWA-MEM. The data quality control steps are summarized in Supplementary Figure 2.

**Table 1:** Datasets used in this study.

| Dataset | Dataset | Status | Number<br>(males/females) | Age in years<br>(SD) |
| --- | --- | --- | --- | --- |
| PROFILE | discovery | Case | 381/126 | 70.8 (8.5) |
| PGPC | discovery | Control | 16/16 | 52.4 (18.5) |
| GTEx | discovery | Control | 97/45 | 50.7 (14.5) |
| TOPMed – IPF | replication | Case | 861/382 | NA |
| TOPMed - CVS | replication | Control | 1364/1833 | NA |
| 1kGP | validation | Control | 50.52 | NA |

*PROFILE - Prospective Study of Fibrosis in the Lung Endpoints,*
*PGPC – Personal Genome Project Canada.*
*TOPMed - Trans-Omics for Precision Medicine*
*1KGP – 1000 Genomes Project phase 3 samples*

### Identification of expanded TRs using Expansionhunter Denovo

TRs showing large expansions associated with IPF risk were identified using PROFILE cases and the GTEX/PGPC controls for discovery and TOPMed case-control dataset for replication. These were identified using ExpansionHunter Denovo (EHdn) software, which identifies TR expansions longer than the sequencing read length that are not necessarily in the GangSTR catalogue and may be rare or de novo expansions ^29^. Individuals were identified through the GWAS quality control protocol described above, and case-control locus-based analysis was performed using EHdn, where anchored in-repeat-reads (IRRs) or both anchored and fully IRRs with the same motif are analysed for each locus. The anchored IRRs are used to estimate the approximate location of the expanded TR, and normalised IRR sequence coverage is used as a proxy of TR expansion size. Expanded loci in cases are identified by comparing normalised IRR counts in cases against controls using a Wilcoxon rank-sum test. Both raw and Bonferroni-corrected (adjusting for multiple tests) *p* values are calculated to determine the significance of expansions.

EHdn does not estimate the allele length but instead reports the approximate location of the expanded repeat to within 500 bp accuracy, so each significant locus was validated by visualisation in the UCSC genome browser against the human reference genome to obtain the repeat genomic coordinates and by using Geneious software (Dotmatics, Boston, USA). To limit the analysis to higher confidence repeats and to avoid false positives we assessed sequence coverage across each TR and at least ±500 bp upstream and downstream. We selected for further analysis TR expansions that showed a significant difference in mean length between cases and controls, with a minimum read depth of 10 and no significant difference in sequence coverage between the two groups. For discovery, a p-value of < 5x10^-8^ was regarded as significant at a genome wide level and a Bonferroni-adjusted p-value of <0.01 was used for replication.

Selected tandem repeats were also re-genotyped using ExpansionHunter v.5.0.0 and GangSTR following the default parameters to obtain individual genotypes, if possible. ^30^

### Genotyping of TR loci using GangSTR

GangSTR was used to genotype STRs (2-6 bp) and VNTRs (7-20 bp) genome-wide in both datasets using a catalogue containing 832,380 TR loci from the human reference genome ^31^.

Variant files generated by GangSTR were filtered using the DumpSTR tool as recommended ^31^, using the following parameters: ## dumpSTR --vcf vcffile --out prefix --vcftype gangstr --gangstr-filter-spanbound-only --gangstr-filter-badCI --gangstr-min-call-DP 20 --gangstr-max-call-DP 1000 --gangstr-min-call-Q 0.9. These parameters were used to remove calls with the maximum likelihood genotype estimates that are outside of the 95% bootstrap confidence interval, calls where only spanning or bounding reads were found, calls with minimum read coverage below 20, calls with abnormally high coverage and calls with a minimum call quality score of below 0.9. The minimum read coverage was set to ensure there is were sufficient sequence reads for credible TR genotype calls. These TR genotype calls were the input for GWAS quality control procedures.

### Genotype quality control

We filtered TR genotype calls by adapting standard approaches for case-control genome wide association studies (GWASs) in order to remove samples and TR loci of poor genotyping quality. Firstly, pairwise linkage disequilibrium between multiallelic TR loci (r̄ ^2^) with unknown phases was determined using well-established methods ^32–34^ implemented in the Pegas R package ^35^. The first two principal components of a subset of TR genotype calls with low pairwise linkage disequilibrium (r̄ ^2^ <0.8) were used to identify and retain individuals of European ancestry, as indicated by clustering with European ancestry samples from the co-analysed 1KGP (Supplementary Figures 3,4). The genetic ancestry identification was cross validated using SNV genotype calls.

TR loci with missingness >1%, individuals with missingness >5%, then related individuals with up to a second degree relative and with sex mismatches were removed. Sex prediction used statSTR software to estimate heterozygosity and assessed the proportion of heterozygous loci for X chromosome calls ^36^. Relatedness was estimated using the KING method implemented in the reletedness2 function in VCFtools ^37,38^. Common polymorphic TR loci were defined as TR loci with at least one alternate allele frequency of >0.01. The same quality control procedures applied to the discovery dataset were also used for selecting individuals identifying loci for replication and further follow-up (Supplementary Figure 5).

### Association testing of common polymorphic TR loci

GangSTR diploid allele length (in repeat units) from each TR locus was analysed assuming a linear relationship between the number of repeats and log-odds of the disease i.e. each increase or decrease in repeat unit has a proportional impact on the odds of the outcome. Two models were fitted: a dominant model which used the number of repeats of the longest allele as the genotype at each locus from each individual, assuming that having only one allele confers the risk of disease and an additive model which analysed total repeat burden at each locus by summing repeat copy number of both alleles.

The following logistic regression model was used to test for association, accounting for age, sex and the first 10 principal components of TR genetic variation: *logit(P(Y=1)) = β_0_+β_1_×STR_i_ + β_2_×covariate_1_ +⋯+β_n_× covariate_n_ ;* β_1_ – effect size of the unit changes in TR length at locus_i_ on the case-control outcome status. *β_2_,…,β_n_ are the effect sizes (coefficients)* for the covariates.

*logit(P(Y=1))* is the log odds of having the disease, STR_i_ represents a continuous variable of an individual’s allele lengths in repeat units for each locus; the longest allele for the dominant model and the sum of two alleles for the additive model. For TOPMed replication dataset age was not included as this information was unavailable.

Statistically significant variants associated with IPF susceptibility were defined as those that met the conventional genome-wide significance threshold (*p* < 5 x 10^−8^). All significant loci were further filtered for departures from Hardy-Weinberg equilibrium (*p* < 1×10⁻⁶) among controls using statSTR software ^36^ . Sequence read alignments for each statistically-significant locus were manually inspected using both the short-read whole genome sequences and high-coverage (∼37x) long-read whole genome sequences from selected individuals of European ancestry from the 1000 Genomes project ^39^. The TR genotype calls at each significant locus was further validated using the alternative TR genotype calling software ExpansionHunter ^30^.

A locus was considered replicated if it met the conventional genome-wide significant threshold (*p* < 5 x 10^-8^) in the discovery phase and at least a Bonferroni-corrected threshold *p* < 0.01 in the replication dataset, with the same direction of effect.

### Conditional Analysis of TR and SNV Effects at known IPF Risk Loci

To assess whether the causal variant at a locus may be the tandem repeat (TR) itself, or a previously reported single nucleotide variant in LD with the TR, we performed conditional regression analyses for TR loci that overlapped (i.e., implicated same gene as the SNV or within 500 kb) with known GWAS SNV signals adjusting for age, sex, and the first ten principal components. For each such locus, we included both the TR genotype and the lead SNV genotype as covariates in a logistic regression model. Each SNV locus was analysed using an additive genetic model with SNVs recorded as dosages (0, 1 or 2), following established methods ^5^.

### Variant functional annotation

IPF-associated TR loci were annotated with putative functional consequences using Annovar software against the RefSeq database and using the Ensembl Variant Effect predictor ^40,41^. Each associated tandem repeat locus was visualised in the UCSC Genome Browser on the human reference genome to obtain the RefSeq sequence ^42^. Genes near associated TR loci were assessed for overlap with previous GWAS signals by checking if the genes had been previously linked to IPF susceptibility or disease progression in the Open Targets platform https://genetics.opentargets.org. Gene function, family and biological pathways were inferred using GENEONTOLOGY https://geneontology.org/. Gene expression data in TPM (transcripts per million) for lung tissues, fibroblasts and lymphoblastoid cell lines were obtained for 362 GTEX samples of European ancestry, and from 360 lymphoblastoid cell lines from the 1000 Genomes Project collection (TSI, n=91, GBR, n=86, CEU, n=91 and FIN, n=92) from Geuvadis https://ftp.1000genomes.ebi.ac.uk/vol1/ftp/data_collections/geuvadis/working/geuvadis_top med. All chromosomal references and analyses refer to the GRCh38 human genome assembly. Statistical analysis was conducted using the statistical language R.

### PCR and Sanger sequencing

Tandem repeats were amplified using standard PCR amplification conditions (Supplementary Table 1), and individual alleles separated based on length by standard gel electrophoresis and gel extraction (Monarch gel extraction kit, New England Biolabs), followed by Sanger sequencing (Source Biosystems) and manual quality inspection. Haploid genomes from the Human Pangenome reference were interrogated using in silico PCR with the PCR primers used above. DNA sequences were analysed for tandem repeat length and sequence variation using Tandem Repeat Finder ^43^.

### Cell culture maintenance and treatments

U-937 (CRL-1593.2 ™) monocyte cells were grown in suspension in RPMI-1640 media supplemented with 10% (v/v) fetal bovine serum, 2 mM L-glutamine, and 100 μg/ml penicillin/streptomycin. Cells were maintained in humified atmosphere with 5% CO_2_ and 95% air at 37°C in a ThermoForma Scientific incubator (Chemie, Valenzano, BA, Italy). Propagation occurred every 2–4 days (70–80% confluence). Detachment of adherent cells was obtained by using 0.5–1 ml of phosphate buffered saline (PBS) containing 0.3% trypsin, 2 mM EGTA, and 0.1% glucose for 1–5 minutes. The action of trypsin was blocked by adding an appropriate volume of culture medium (containing α2-antitrypsin) to the cell suspensions. All experiments with the cell lines were performed between passages 3 and 6 of post-thaw propagation.

Cultured cells were used in NMD system inhibition experiments by administering cycloheximide (CHX) dissolved in DMSO (dimethyl sulfoxide) to cells cultured at 90% confluence (final concentration of 100 µg/mL CHX in fresh culture medium) for 1, 2, and 4 hours. The untreated control was administered with fresh culture medium for 4 hours, supplemented with a volume of DMSO equal to the inhibitor volume added for treatments.

### Total RNA extraction from cell cultures

RNA extractions from cells were performed using the All-Prep DNA/RNA/Protein mini kit (Qiagen, Hilden, Germany) according to the manufacturer’s instructions. Cultured cells were double-washed with D-PBS and then lysed with the kit lysis buffer by scraping. RNA concentrations were calculated by a NanoDrop ND-2000 Spectrophotometer (Nanodrop Technologies, Wilmington, DE, USA), and RNA samples were tested qualitatively by electrophoresis on 1% (*w*/*v*) agarose gels.

### Reverse-transcriptase PCR (RT-PCR)

Details of the oligonucleotide sequences are reported in Supplementary Table 1. The SuperScript® III RT kit (Thermo Fisher Scientific, Monza, Italy) was used to perform reverse transcription reactions with 500 ng total RNA extracted from cell lysates. The Platinum®Taq DNA Polymerase (ThermoFisher) was used for subsequent PCR experiments. As an internal control, primer pairs specific for the 18S ribosomal RNA were added to the mixture together with the gene-specific primers.

For RT-PCR conducted under semiquantitative conditions (relative quantitative), the Ambion QuantumRNA Classic II 18S Internal Standard kit (Thermo Fisher Scientific, Monza, Italy) was used. In this case, random hexamers were used for reverse transcription, and the reverse transcription mixture was pre-incubated for 5 minutes at 25°C, according to the protocols for the SuperScript III RT kit. For the PCR reaction, primers and competimers for 18S ribosomal RNA (expected PCR product of 324 bp) were added to the reaction mix according to the Ambion kit protocol. For the gene under study, a 18S primer:competimer ratio of 1:9 was selected (the ratio at which the gene-specific and 18S amplification bands were densitometrically comparable). The optimal number of amplification cycles for the gene-specific primer pair was chosen based on the amplification linearity range, assessed by amplifying ten different aliquots of the same reaction mixture for an increasing number of cycles (from 21 to 39). Densitometric analysis of the intensity of the bands from the ten amplifications, loaded on the same agarose gel, allowed the aforementioned range to be calculated. For the semiquantitative analysis of mRNA expression, a number of cycles within the linearity range was chosen, that is 35 cycles for the human *SLC15A4* transcripts. Each semiquantitative RT-PCR experiment was repeated twice; the values reported in the experimental results are the average of the normalized values obtained from three independent biological replicates.

## Results

We used a case-control design for discovery, with a dataset consisting of 507 cases and 174 controls, and the replication dataset consisting of 1243 cases and 3197 controls, all whole genome sequenced using Illumina paired-end read sequence to a sample average of at least 30X coverage. We then conducted a genome-wide association study using TRs called by two complementary techniques: ExpansionHunter denovo (EHdn) or GangSTR. EHdn compares the number of anchored sequence reads (i.e., a sequence read which partially maps to sequence flanking the TR) to sequence reads that map within the repeat to determine whether there is an expanded allele in certain individuals ^29^. Importantly, EHdn is tolerant of complex repeats and does not need a catalogue, so it can detect rarer expansions at novel TRs. GangSTR uses a catalogue of known TRs to estimate actual sample genotypes at each TR by integrating several pieces of information for each TR, such as coverage and fully-spanning reads ^31^. An overview of the experimental strategy is shown in Figure 1.

**Figure 1 –.**
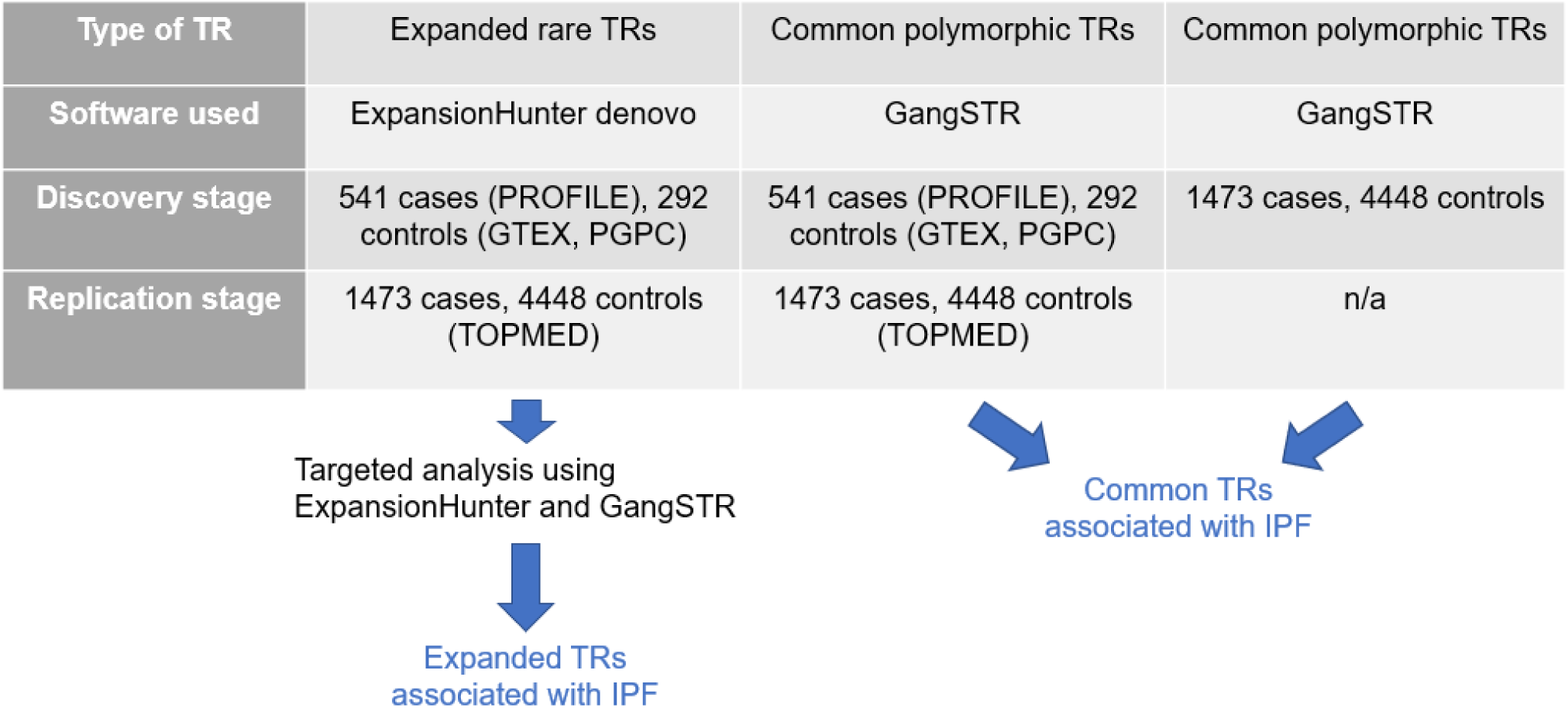
Experimental design and analysis strategy.

### Tandem repeat expansions associated with IPF

We initially asked whether IPF was associated with any particular expansion of a TR across the genome, using the software EHdn. Ten repeat expansions were identified in the discovery phase, of which three were replicated using the TOPMed dataset (Table 2, Supplementary Table 2). Of the three significant loci, one affected an exon of the non-coding RNA gene *LINC03021*, one was in intron 2 of the histidine-transporter gene Solute Carrier Family 15 member 4 (*SLC15A4*) affecting a likely enhancer element as identified by GeneHancer, and the third locus was 16.9kb distal of the G-Patch Domain Containing 2 Like (*GPATCH2L)* gene.

**Table 2:**
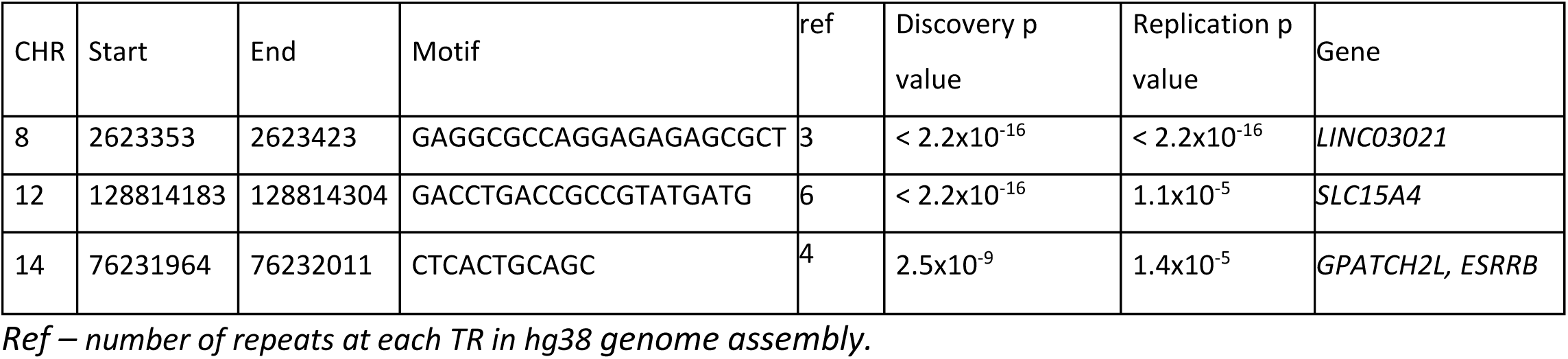
TR loci with expansions associated with IPF susceptibility.

### Common tandem repeats associated with IPF

We asked whether IPF was associated with common TRs, genotyped using GangSTR. A total of 684,651 autosomal TRs were genotyped in more than 90% of the individuals, of which 96,181 were polymorphic (Supplementary Figure 1). Following GWAS quality control to select high-quality TR genotypes and individuals of European ancestry, 507 IPF cases, 181 controls and 25,144 TR autosomal loci (call-rate ≥ 99%) were available for discovery analysis (Table 3).

**Table 3:** Genome-wide TR loci associated with IPF susceptibility.

| CHR | Start | End | Motif | Ref | Rs ID | Discovery<br>Odds<br>Ratio<br>(95% CI) | Discovery<br>P value | Discovery<br>N alleles | Replication<br>Odds Ratio<br>(95% CI) | Replication<br>P value | Replication<br>N alleles | Gene |
| --- | --- | --- | --- | --- | --- | --- | --- | --- | --- | --- | --- | --- |
| 4 | 9712251 | 9712270 | TATT | 5 | rs58143886 | 0.22<br>(0.12-0.33) | 6.1x10<sup>-9</sup> | 3 | 0.58 (0.5-0.66) | 8.8x10<sup>-16</sup> | 3 | *ENSG00000301897* |
| 9 | 63866153 | 63866176 | AAAT | 6 | rs560545907 | 0.25<br>(0.15-0.9) | 4.7x10<sup>-10</sup> | 4 | 0.9 (0.73-1.1) | 0.31 | 4 | *FRG1JP -*<br>*FLJ43315* |
| 20 | 30629426 | 30629440 | GCTGT | 3 | rs1228828566 | 0.02<br>(0.01-0.06) | 8.2x10<sup>-11</sup> | 2 | 0.34 (0.12-0.76) | 0.016 | 2 | *FRG1BP - DEFB115* |

CI – confidence interval, Ref –number of repeats at each TR in the reference genome. N alleles – total number of different allele
types at each locus among cases and controls analysed.

Under an additive model, with genomic inflation well-controlled (Supplementary Figure 6), three TR loci were identified as associated with disease risk in the discovery case-control dataset (Table 3). One TR was between the *FRG1JP and FLJ43315* genes, one TR was in intron 2 of a novel long non-coding transcript *ENSG00000301897* (∼70 kb from the *DRD5* gene) and the third locus was a TR located near the non-coding RNA *LOC102723769* between the pseudogene *FRG1BP* and the beta-defensin gene *DEFB115*. These three loci were genotyped by GangSTR and ExpansionHunter in a total of 1243 cases and 3197 controls individuals of European ancestry obtained from TOPMed for replication (Table 1) and only the association in *ENSG00000301897* was replicated (Table 3), although the other two loci showed genotypes not in HWE in the replication dataset, suggesting inaccurate genotyping of these loci in that dataset. GangSTR allele frequencies for these loci matched those of ExpansionHunter for both discovery and the replication dataset.

Because of the age disparity between the PROFILE cases and the GTEX controls analysed in the discovery dataset (Table 1), we assessed the impact of age by including all covariates from the initial analysis except age in a further regression analysis. A slight shift in significance level was observed; however, no additional associations reached genome-wide significance and replicated.

### Tandem repeat GWAS for IPF using the TOPMed dataset

Given the limited power of the initial discovery analysis to find common TRs of small effect size, we also conducted a discovery TR GWAS using the large TOPMed dataset as cases and controls alone. A total of 51,766 loci (with minimum locus call rate >= 99%) were genotyped using GangSTR in 1,243 cases and 3,197 controls. After genome-wide association analysis, with genomic inflation well-controlled (Supplementary Figure 6), eight significant TR loci were identified, one at the 11p15.5 locus (near the *MUC5AC* and *MUC5B* genes), the remaining seven clustered at the 17q21.31 locus (near the *KANSL1* and *MAPT* genes) (Figure 2).

**Figure 2 –.**
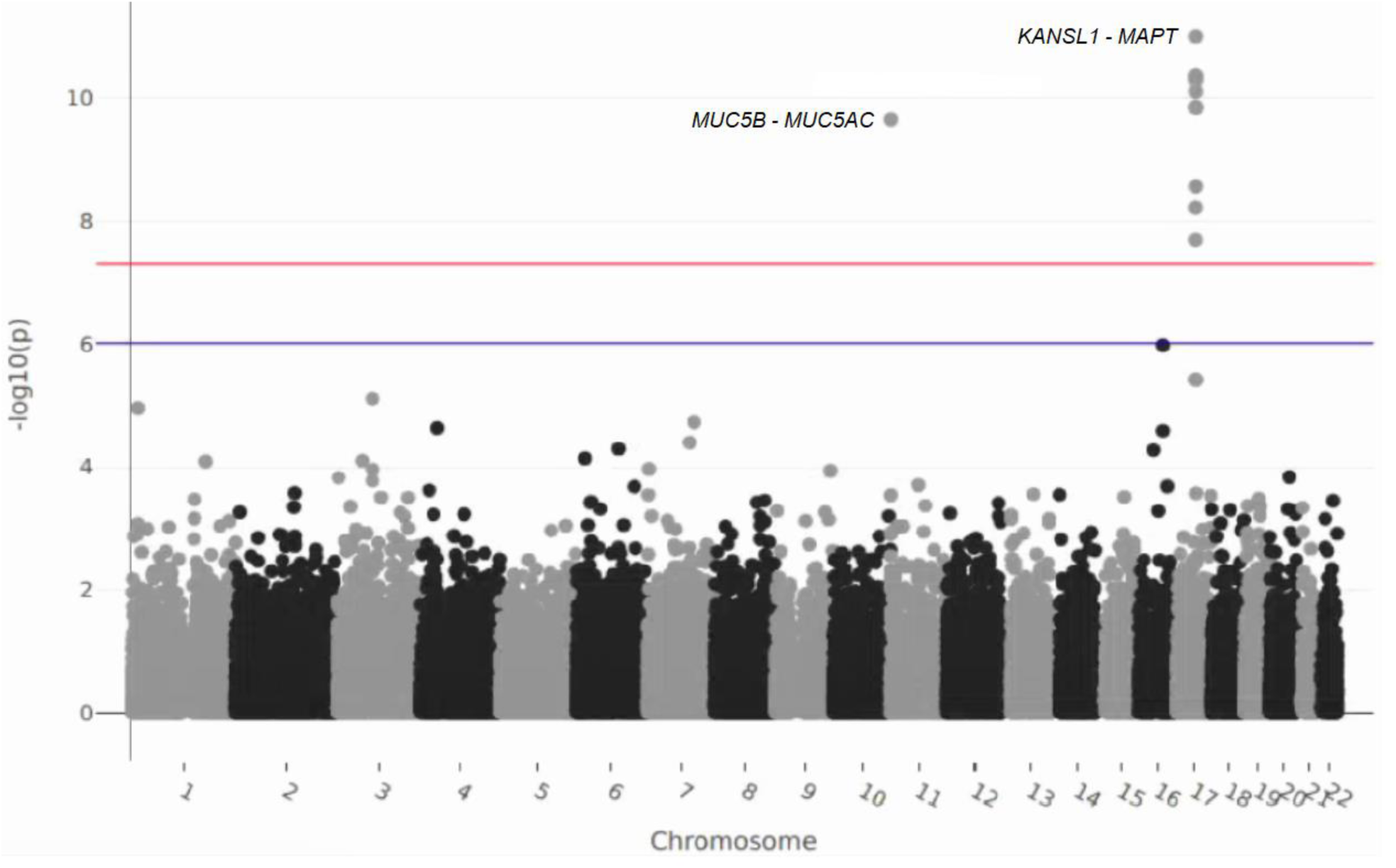
Association of TRs with IPF in the TOPMed dataset. *Association data for 1243 cases and 3197 controls genotyped for 51766 loci using GangSTR, with data points coloured black/grey according to alternating chromosomes, p=1x10^-6^ shown as a blue line, and p=5x10^-8^ shown as a red line*.

Both loci have been previously associated with IPF in GWAS, which provided us with reassurance that this TR-GWAS was technically accurate enough, and had power to, detect previous associations through LD with previously-associated SNVs. We tested whether the signals from the SNV and the TR reflect the same underlying association at the 17q21.31 locus and at the 11p15.5 locus, using genotype data from TOPMed, with models including both variant types to assess their independent effects. At the 11p15.5 locus (*MUC5B*), the TR lost genome-wide significance (*p*=xxx) when the promoter SNV rs35705950 was included, indicating that the SNV and the TR represent the same association signal. The SNV remained highly significant regardless of TR inclusion (OR = 4.4, *p* = 2.1 × 10⁻⁸⁵ with TR; OR = 4.5, *p* = 1.4 × 10⁻⁹⁴ without TR) suggesting that the SNV represents the actual functional variant. At the 17q21.31 locus, neither the lead SNV (rs2077551) nor any of the three TRs mapping to 17q21.31 remained significant when modelled together suggesting that all reflect the same underlying functional variant.

### Internal structure of SLC15A4 tandem repeats

Any functional effect of tandem repeats might be due to length variation or sequence variation between repeat units. Because it disrupted an annotated enhancer element within a gene, we decided to focus on the *SLC15A4* TR. We investigated the sequence variation between the repeat units, including the SNV rs3741615, immediately distal to the last nucleotide of the *SLC15A4* TR, their relationship with *SLC15A4* TR length and disease association. We examined the sequence of the gold standard genotyped TR alleles described above from the haploid genome assemblies from the Human Pangenome Consortium, and also directly PCR amplified and Sanger sequenced from CEU samples. The *SLC15A4* TR exhibited extensive sequence variation between individual repeat units. Each repeat unit distinguished by sequence was assigned a letter, with the common reference repeat designated A, and other letters representing repeat units that differ by a single nucleotide positions (Figure 3a).

**Figure 3.**
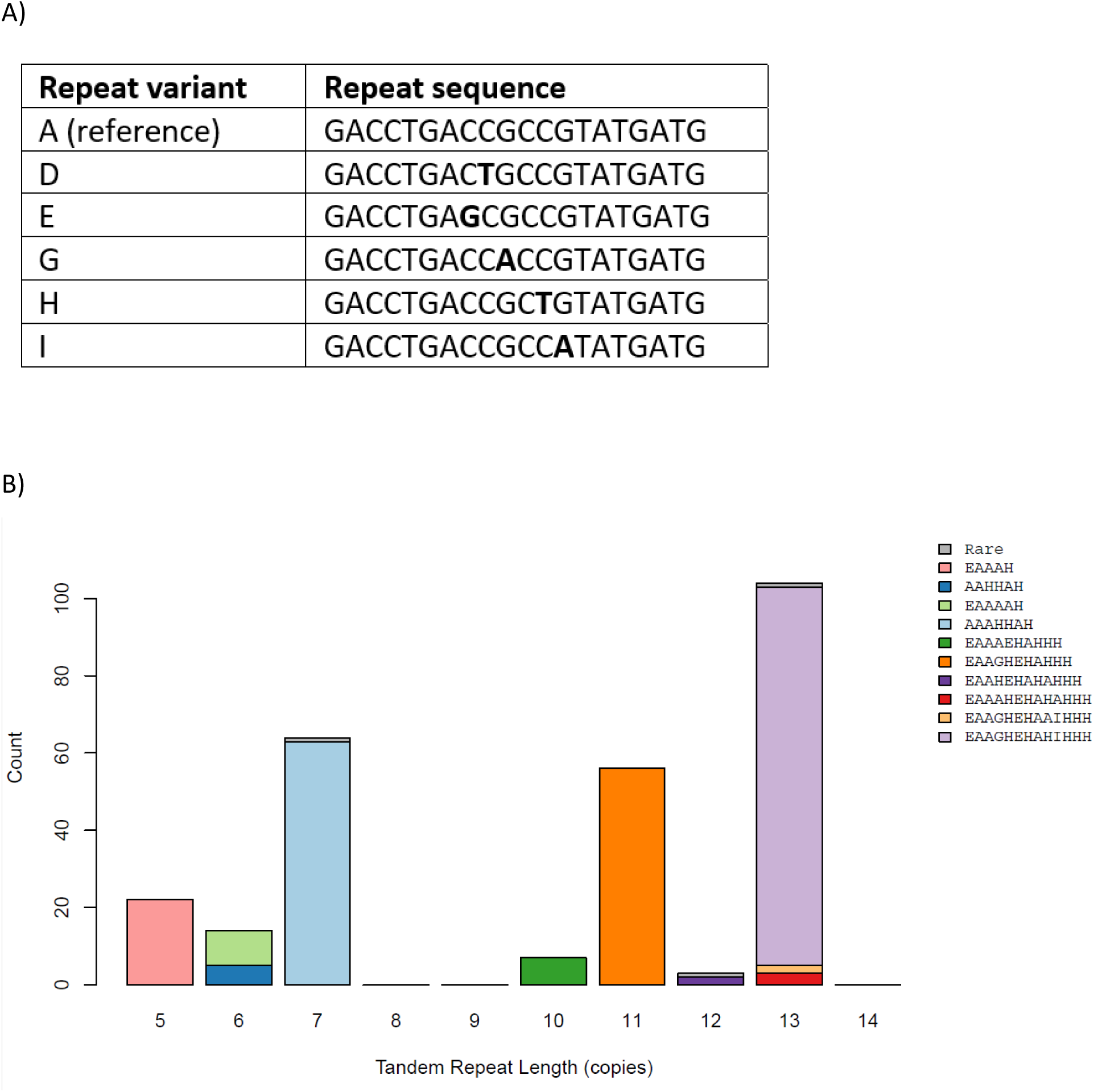
Sequence variation within *SLC15A4* tandem repeats. A) Key showing the repeat variant code and the 20bp repeat variant sequence, with the sequence differences from the reference sequence (variant A) highlighted in bold. B) The barplot is coloured according to the sequence composition of the length allele, as shown by the key on the right. Sequence alleles that occur only once are labelled in grey as “rare”. n=270.

The *SLC15A4* tandem repeat has seven length alleles ranging from 5 to 13 tandem repeats, bimodally distributed into long (>9) and short alleles (Figure 3b). There is extensive variation of repeat sequences (Figure 3a), which assort into 13 alleles distinguished by length and sequence variation from a total of 270 chromosomes analysed. Alleles of a particular length are almost always identical in terms of sequence structure, with the exception of the 6 copy allele, where two different sequence structures are common.

Longer alleles are characterized by an expansion of the tandem repeat sequence H at the distal end of the repeat, making the longer alleles somewhat distinct from shorter alleles not only by number of tandem repeats but also by sequence structure.

We used our data to investigate the association between rs3741615 and the TR alleles. The rs3741615-A allele was exclusively associated with the 5 copy allele at the TR, and this is shown when comparing the length of alleles on rs3741615-A versus rs3741615-G backgrounds (t = -23.62, p = 1.95 × 10⁻⁶⁵), with the rs3741615-G allele associated with longer TR alleles. This association suggests that the sQTL and eQTL effects detected for rs3741615 may in fact reflect effects caused by the TR.

Given the strong association between the TR and rs3741615, we asked whether any association with IPF is detectable in previous SNV-based GWAS studies. Using data from the previous IPF GWAS meta-analysis ^3^, we found that the rs3741615-G allele was associated with an increased risk of IPF, albeit not at genome-wide significance levels (*p*=1.45x10^-3^, OR=1.17, 95%CI 1.06-1.28), and the direction of effect was consistent in four of the five studies analysed.

### Expression effect of tandem repeats

We investigated how the TRs associated with IPF might exert their effect on disease risk. Inspection of human genome annotation ^42^ shows that the *LINC03021* TR is within an ENCODE candidate cis-regulatory element, and the *SLC15A4* TR is flanked by candidate cis-regulatory elements and is within a candidate cis-regulatory element. Because these two TRs disrupt regulatory elements, it seemed plausible that these TRs are eQTLs for a nearby gene. We therefore used the GTEx RNAseq data to measure the level of expression of the nearest genes for each TR in three biologically-relevant cells/tissues: fibroblasts, lung tissue and lymphoblastoid cells. We then correlated tandem repeat length against transcript level using tandem repeat genotypes called by ExpansionHunter on the GTEx/Geuvadis dataset, on samples with a genome sequence coverage >30x.

*SLC15A4* expression (Figure 4a,4c) was negatively correlated with TR length in fibroblasts (*p*=0.025). No association was found between TR copy number and expression of *GPATCH2L* or *DRD5,* and *LINC03021* was not expressed in the three tissues analysed. The single nucleotide variant immediately distal to the last nucleotide of the *SLC15A4* TR (rs3741615), and affecting the same annotated regulatory element, was an eQTL for *SLC14A5* in fibroblasts (*p*=4.8x10^-15^) with the rs3741615-A allele associated with higher expression.

**Figure 4 –.**
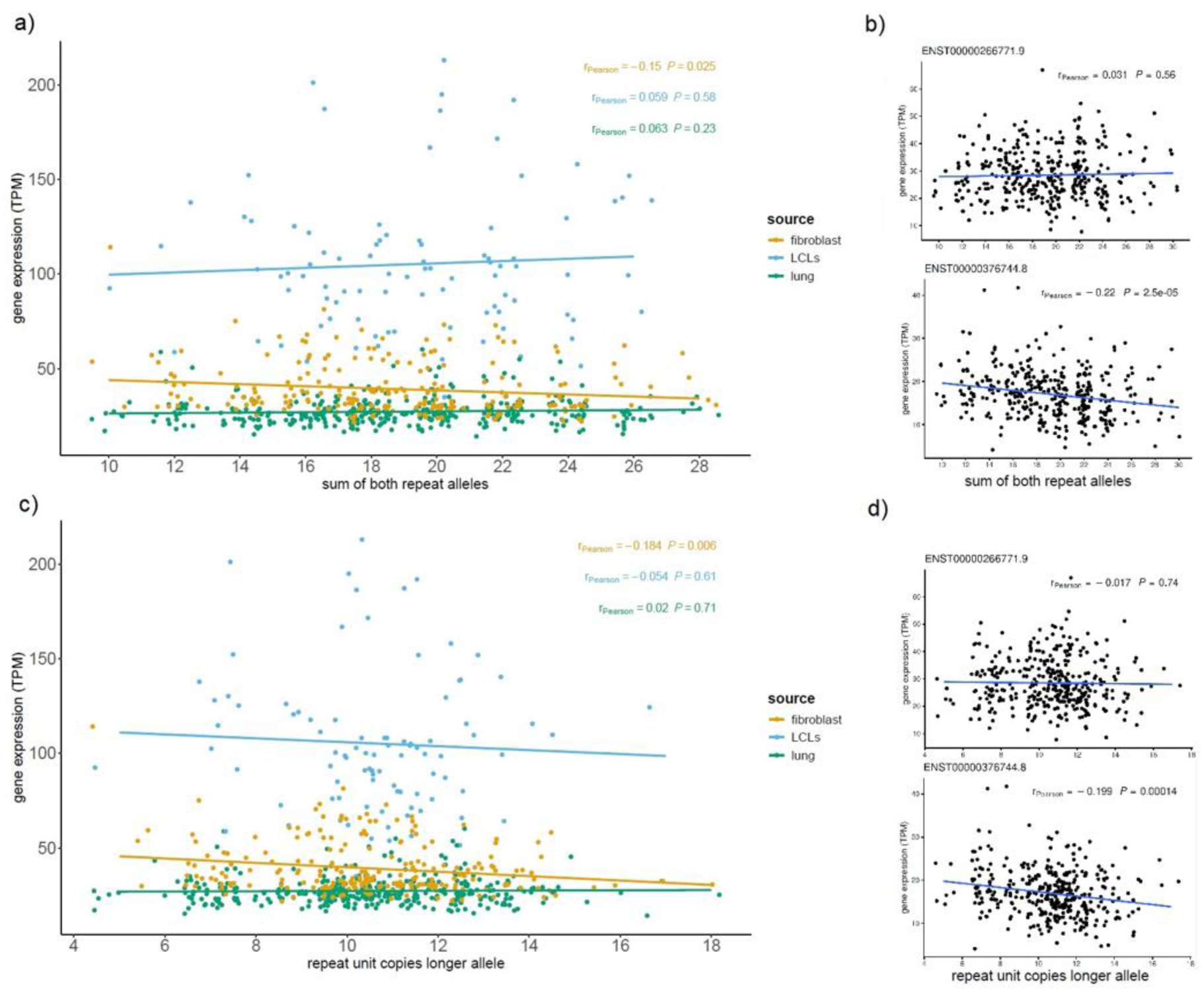
Correlation of *SLC15A4* gene expression with TR length. *a) Correlation of gene expression levels with tandem repeat length, displayed as the sum of both alleles. Transcript data are from GTEx for three cell/ tissues: fibroblast (yellow, n=221), lymphoblastoid cells (blue, n=90) and lung (green, n=361). Horizontal jitter is added in plotting to each repeat allele to enhance the visibility of individual datapoints.* *b) Correlation of transcript expression levels in lymphoblastoid cells with tandem repeat length, displayed as the sum of both alleles (n=360).* *c)-d) As a)-b), except only the longest tandem repeat per genotype is analysed (dominant model)*.

Analysis of GTEx data ^44^ showed that rs374165 was also a splicing QTL for introns of *SLC15A4* in several relevant tissues (Table 3), in lung (*p*=4.6x10^-11^), and in lymphocytes (*p*=1.2x10^-10^), with the rs374165-A allele associated with increased amounts of splicing involving exons 7 and 8. Furthermore, for lung and fibroblasts, the rs374165-A allele was associated with decreased amounts of use of a cryptic splice site within intron 7 (chr12: 128794356). When used, this cryptic splice site would generate a transcript with a premature stop codon and, if translated, a SLC15A4 protein lacking the final C-terminal transmembrane domain. This splice site is also used by transcript variant X3 (XR_007063044.1), a non-coding transcript. The rs3741615-A allele is also more weakly associated with alternative splicing in fibroblasts resulting in skipping of exon 3 in the final transcript, resulting in the truncated transcript version X6.

Because of the potential effects of the tandem repeat on splicing, we decided to analyse the association between the TR, genotyped by ExpansionHunter, and the expression levels of specific transcripts, using the Geovadis lymphoblastoid RNASeq dataset on the 1000 Genomes project samples. Of the eight transcripts analysed, one transcript (ENST00000376744.8) showed a significant decrease in expression associated with increasing tandem repeat length (additive model, Pearson’s r=-0.22, *p*=2.5x10^-5^, Figure 4b; dominant model, Pearson’s r=-0.199, *p*=1.4x10^-4^, Figure 4d). This transcript excluding exon 3 corresponds to the NCBI-annotated isoform X6 (Table 4). Expression levels of the full-length canonical transcript (ENST00000266771.9) showed no correlation with tandem repeat length (Figures 3b, 3d), suggesting that the tandem repeat exerts its functional effect through levels of a short *SLC15A4* transcript isoform.

**Table 4.**
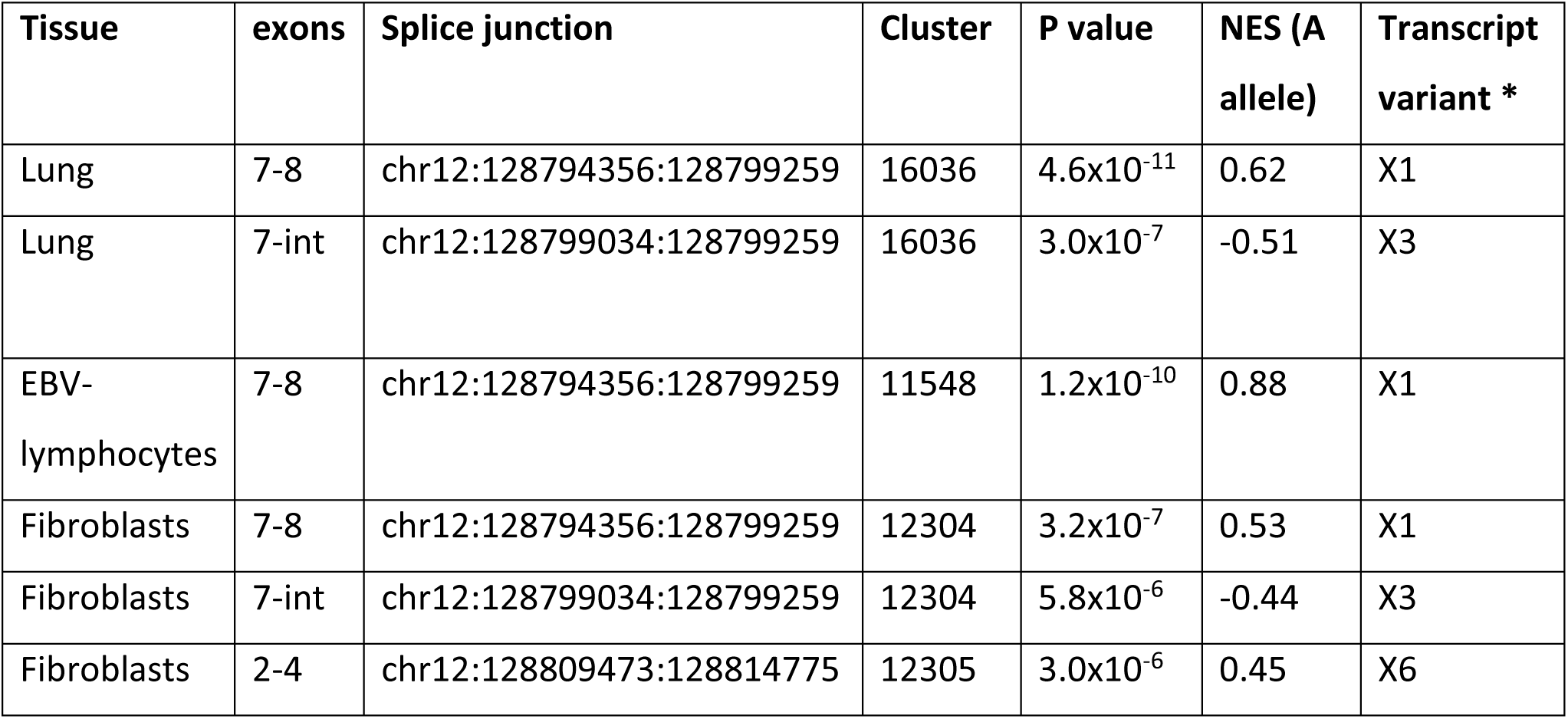
Analysis of rs3741615 sQTL. * Transcript variant indicates the annotated RefSeq transcripts that use that splice junction

| Tissue | exons | Splice junction | Cluster | P value | NES (A allele) | Transcript variant * |
| --- | --- | --- | --- | --- | --- | --- |
| Lung | 7-8 | chr12:128794356:128799259 | 16036 | $4.6 \times 10^{-11}$ | 0.62 | X1 |
| Lung | 7-int | chr12:128799034:128799259 | 16036 | $3.0 \times 10^{-7}$ | -0.51 | X3 |
| EBV-lymphocytes | 7-8 | chr12:128794356:128799259 | 11548 | $1.2 \times 10^{-10}$ | 0.88 | X1 |
| Fibroblasts | 7-8 | chr12:128794356:128799259 | 12304 | $3.2 \times 10^{-7}$ | 0.53 | X1 |
| Fibroblasts | 7-int | chr12:128799034:128799259 | 12304 | $5.8 \times 10^{-6}$ | -0.44 | X3 |
| Fibroblasts | 2-4 | chr12:128809473:128814775 | 12305 | $3.0 \times 10^{-6}$ | 0.45 | X6 |

### Short SLC15A4 transcript isoforms are targeted by nonsense-mediated mRNA decay

Using the human monocyte-like cell line U937, we directly detected *SLC15A4* transcripts with alternative splicing patterns involving intron 2 and exon 3. In particular, two different forms of *SLC15A4*-related transcripts show the presence of a shorter exon 3 (transcript variant X5) or the total absence (skipping) of exon 3 (transcript variant X6, Table 4) respectively, compared to the full-length canonical transcript. Comparison with the canonical reference nucleotide sequence reveals that the X5 sequence contains a gap corresponding to the absence of the upstream 58-nucleotide sequence of the canonical exon 3, while the X6 sequence shows a gap of 168 nucleotides corresponding to the lack of the entire exon 3 (Figure 5a).

**Figure 5.**
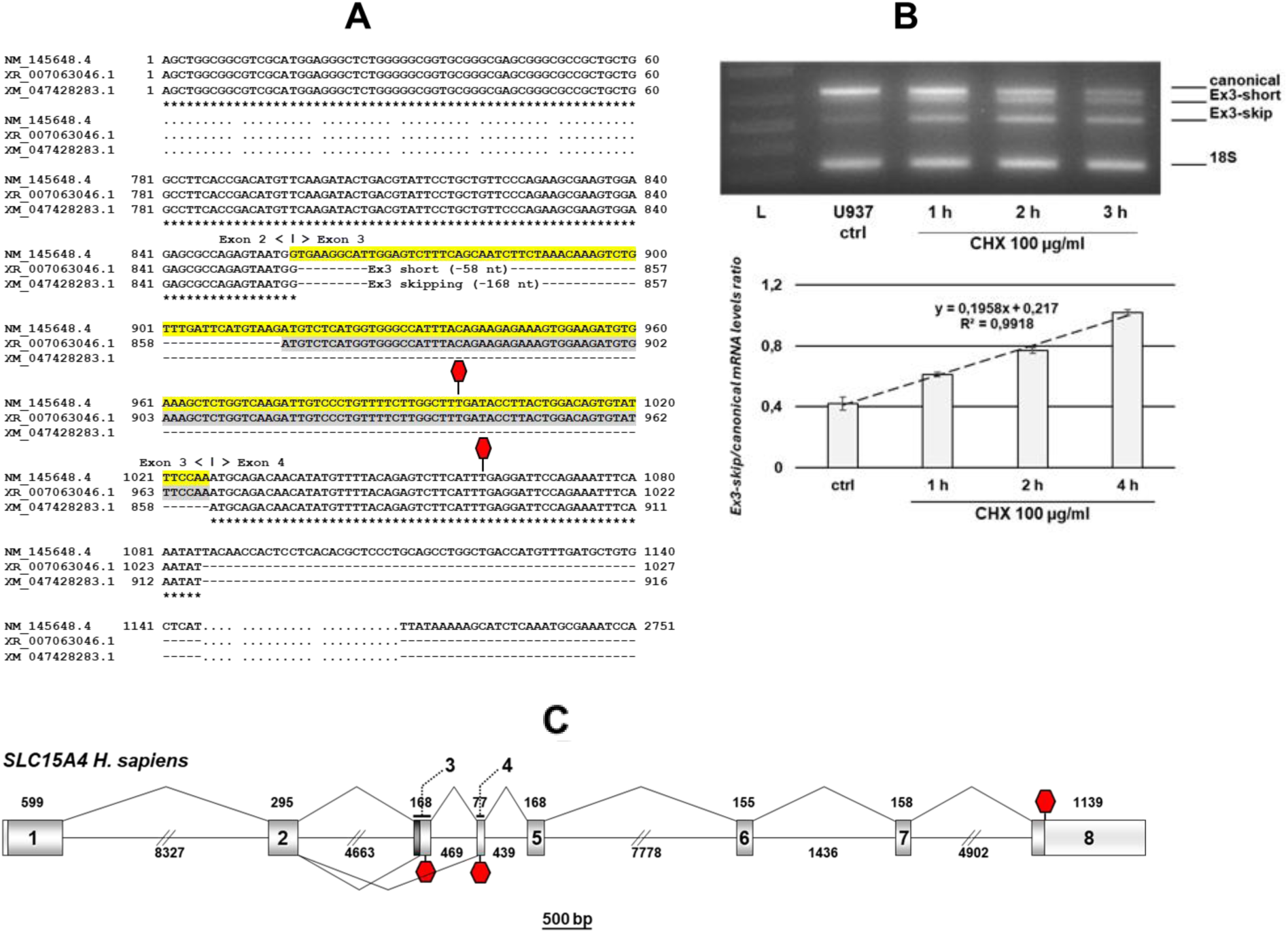
Analysis of *SLC15A4* transcripts under inhibition of nonsense-mediated decay. **(A)** Multiple sequence of transcript variant X5 (XR_007063046.1), transcript variant X6 (XM_047428283.1) and canonical (NM_145648.4) mRNA sequences. Exon 3 sequence is highlighted in yellow in the canonical *SLC15A4* mRNA. Transcript variant X5 shows a 58 nt gap, indicating the alternative short exon 3 (Ex3 short); the 168 nt gap is in transcript variant X6 (XM_047428283.1), indicating the alternative skipping of exon 3 (Ex3 skip). **(B)** RT-PCR of *SLC15A4* mRNAs in total RNA extracted from U397 cells in NMD system inhibition experiments with 100 µg/ml CHX. The sample shows the amplification product of 18S RNA (housekeeping internal control), the ∼700 bp product corresponding to the human *SLC15A4* mRNA (canonical), the ∼530-bp amplification product (Ex3-skip), and a faint amplification product (Ex3-short) ∼50-60 bp shorter than canonical. Relative levels of constitutive and PTC-containing mRNA isoforms represent densitometric values of the amplification products, normalized to 18S rRNA. Mean +/- SE of 3 independent biological replicates. **C)** Structure of the *SLC15A4* gene with exon and intron sizes. In exons 1 and 8, light grey boxes represent UTR regions. Alternative splicing events are drawn below the gene, while canonical spliced introns are drawn above the gene. Red hexagons represent termination codons.

To detect splicing events involving intron 2 and exon 3, specific primer pairs for RT-PCR assays were designed on the reference *SLC15A4* mRNA sequence, i.e. a sense primer in exon 2 and an antisense primer in exon 5. We detected an amplification product of ∼700 bp, as expected for the canonical human *SLC15A4* mRNA, and a second intense band of ∼530 bp, i.e. a cDNA ∼170 bp shorter (Figure 5b). A third faint signal corresponded to a cDNA lacking ∼50-60 bp relative to the ∼700 bp amplicon. Both detected splicing events affect the coding sequence, resulting in a loss of the reading frame that introduces a premature termination codon (PTC) for protein synthesis. Specifically, skipping of exon 3 in X6 leads to a frameshift that creates a PTC in exon 4, while use of the 3’ alternative splice site generating the short exon 3 in X5 produces a shortened exon 3 that introduces a PTC in exon 3 (Figure 5c). Both PTCs occur more than 50 nucleotides upstream of the last exon-exon junction; this positioning suggests that these alternative transcripts are likely to be targeted for degradation by the NMD (Nonsense-Mediated Decay) surveillance system.

To assess whether the human *SLC15A4* transcript variant X6, showing skipping of exon 3, is indeed a substrate of the NMD system, we evaluated the stability of splice variants in the presence of specific NMD system inhibitors. To block NMD degradation, we used cycloheximide (CHX), a protein synthesis inhibitor that, by blocking translation elongation, effectively prevents PTC recognition (and thus the degradation of PTC-containing transcripts). U937 cells were treated with cycloheximide for 1, 2 or 4 hours, and the expression levels of both canonical *SLC15A4* mRNA and X6 transcript variants were measured by RT-PCR. A time-dependent accumulation of the transcript variant X6 was observed under CHX treatment, together with a time-dependent decrease of the canonical mRNA (Figure 4b). Overall, these results clearly reveal that transcript variant X6 is rapidly recognized and degraded by the NMD system, and that levels of transcript X6 are inversely related to levels of the canonical transcript.

## Discussion

We have conducted a genome-wide association study of tandem repeat variation and idiopathic pulmonary fibrosis susceptibility using two complementary software approaches (GangSTR and EHdn) and whole genome sequences. Two other genotyping approaches (ExpansionHunter and PCR followed by Sanger sequencing) were used to validate the robustness of the positive associations.

Four novel loci were found to be associated with IPF – one within an intron of the gene *SLC15A4*, one within a non-coding RNA, and the other two near the genes *GPATCH2L* and *DRD5*. Because it disrupts an enhancer element within a gene, the most convincing locus is a complex heterogeneous TR within intron 2 of *SLC15A4*, 470bp downstream of the exon 2 splice-site, where expansion of the TR is associated with increased risk of IPF. *SLC15A4* is a gene encoding a 12-membrane-spanning histidine transporter (peptide-histidine transporter 1, PHT1), involved in transporting histidine and certain di- and tri-peptides, is ubiquitously expressed at moderate levels across tissues ^45^, and, within cells, is present in the membrane of endolysosomes ^46^. Knockout mice and deletion studies in human cells show that SLC15A4 functions downstream of the nucleic acid-sensing endosomal Tlr7/9 receptors in the interferon regulatory factor (IRF) pathway (but not MAPK or NF-kB pathways), involved in response to viral infection ^47–51^. This clearly implicates inflammation and dysregulation of the immune response as components of IPF etiology, at least in the early stages when repeated injury to lung alveoli triggers the early development of disease.

Our results show that expansion of the TR within *SLC15A4* may affect the risk of IPF by mediating alternative splicing rather than by controlling overall transcription levels driven by the canonical *SLC15A4* promoter. Longer TR alleles reduce the expression of a short transcript (X6) of *SLC15A4* which is usually degraded by nonsense-mediated decay. The consequences of this are not clear, as in our experimental cell model, but not in our population data, this reduction of the level of the X6 transcript is accompanied by a relative increase in the level of the full-length transcripts, which therefore could potentially increase the amount of SLC15A4 protein. Further experiments should aim to explore the effect of this apparent isoform switch on SLC15A4 protein levels. It is also essential to understand how the length and sequence variation of the tandem repeat controls splice site usage.

Other potential loci were discovered which deserve further investigation. *DRD5* is a dopamine receptor expressed primarily in the brain (Human Protein Atlas, proteinatlas.org, ^45^), but agonists for the closely-related receptor DRD1 shift the phenotype of lung mesenchymal cells from pro-fibrotic to fibrosis-resolving, reversing in vitro extracellular matrix stiffening and in vivo tissue fibrosis in mouse models ^52^. *GPATCH2L*, G-patch-containing 2-like, is expressed at moderate levels ubiquitously ^45^ and has no known function. Mice with homozygous knockout of the close paralogue, *GPATCH2*, are viable and appear normal ^53^, although overexpression and knockdown studies affect cell proliferation and cell division, at least in the 293T human kidney cell line ^54^.

Our study highlights the strengths of studying disease associations with tandem repeats. A key strength is that novel loci will be identified, because some TRs are in low linkage disequilibrium with SNVs, and therefore disease associations will not be reliably detected by flanking SNVs. Furthermore, although low linkage disequilbrium limits the ability of many TRs to be imputed from genomewide SNV genotypes currently, some are in sufficient LD to either reflect associations previously detected as genomewide significant in GWASs (as shown for the TR analysis in the TOPMed dataset), or for GWASs to detect associations at sub-genomewide significance levels due to LD with the causative TR, as shown for the *SLC15A4* locus here ^17^. Both are important validations confirming the robustness of the results.

While short-read whole genome sequences clearly allow successful genome-wide genotyping of TRs ^31,55^, there remain limitations because of the heterogeneity of these repeats including repeat unit length, overall length, and sequence diversity. These limitations vary with the quality of the short-read genome sequence, meaning results from cohorts sequenced and analysed using subtly different approaches are vulnerable to batch effects. The increasing number of whole genomes generated by long-read sequencing will help address at least some of these problems not only by direct genotyping of TRs from these sequences but also by refining alignment-based TR calls from short-read sequencing and developing larger imputation panels to increase the reliability of TR imputation from flanking variation. These approaches are likely to allow the discovery of novel loci involved in many different diseases. A further limitation is under-representation on individuals of non-European ancestries in these studies, which results in under-sampling of allelic diversity and potential limiting the application of findings for non-European individuals.

SLC15A4 recruits the innate immune adaptor TASL and is a critical component of the Tlr7/9 pathway, sensing nucleic acids and triggering the production of interferons and proinflammatory cytokines ^56,57^. Our study suggests that dysregulation of this innate immune pathway, involved in the antiviral inflammatory response ^51,58^, is important in the etiology of IPF and further study will open up novel, and potentially personalized, therapeutic avenues ^59,60^.

## Data Availability

All data produced are available online at https://doi.org/10.25392/leicester.data.26317423.v2

## Acknowledgements

We would like to thank Bruce Weir (University of Washington) for helpful discussions on linkage disequilibrium, and Diana Martin for technical support. JWO is funded by a Wellcome Trust PhD studentship as part of the Wellcome Trust Genetic Epidemiology and Public Health Genomics Doctoral Training Programme by grant number 218505/Z/19/Z. LWV holds a GSK/Asthma+Lung UK Chair in Respiratory Research (C17-1). OS was funded by the Turkish Ministry of National Education, Republic of Turkiye postgraduate study abroad program and a BBSRC Impact Accelerator Award. The research was partially supported by the National Institute for Health Research (NIHR) Leicester Biomedical Research Centre, and the Italian Ministry of Universities and Research (PRIN, Bando 2022 prot. No. 20228XHTBZ). This research used the ALICE High Performance Computing facility at the University of Leicester.

## Author contributions

JHO – Data curation, formal analysis, investigation, visualization, writing-original draft

AA – investigation, validation

RA – Formal analysis

NS - formal analysis

BW and OS – investigation

GS, TM, PM and IS – Resources

AB, AR – investigation, validation, visualization, writing-original draft

TV – investigation, validation, funding acquisition

LVW – Conceptualization, supervision, funding acquisition

EJH – Conceptualization, investigation, supervision, validation, visualization, writing – original draft.

All authors were involved in Writing – review and editing

## Data availability

https://doi.org/10.25392/leicester.data.26317423.v2

## Code availability –

Code for sequence alignment and quality control using Qualimap, TR genotyping using GangSTR, Expansionhunter, and ExpansionHunter Denovo, Phasing and Imputation using Beagle and LD calculation using R is available at https://doi.org/10.25392/leicester.data.26317423.v2

---

**Supplementary Figure 1 –.**
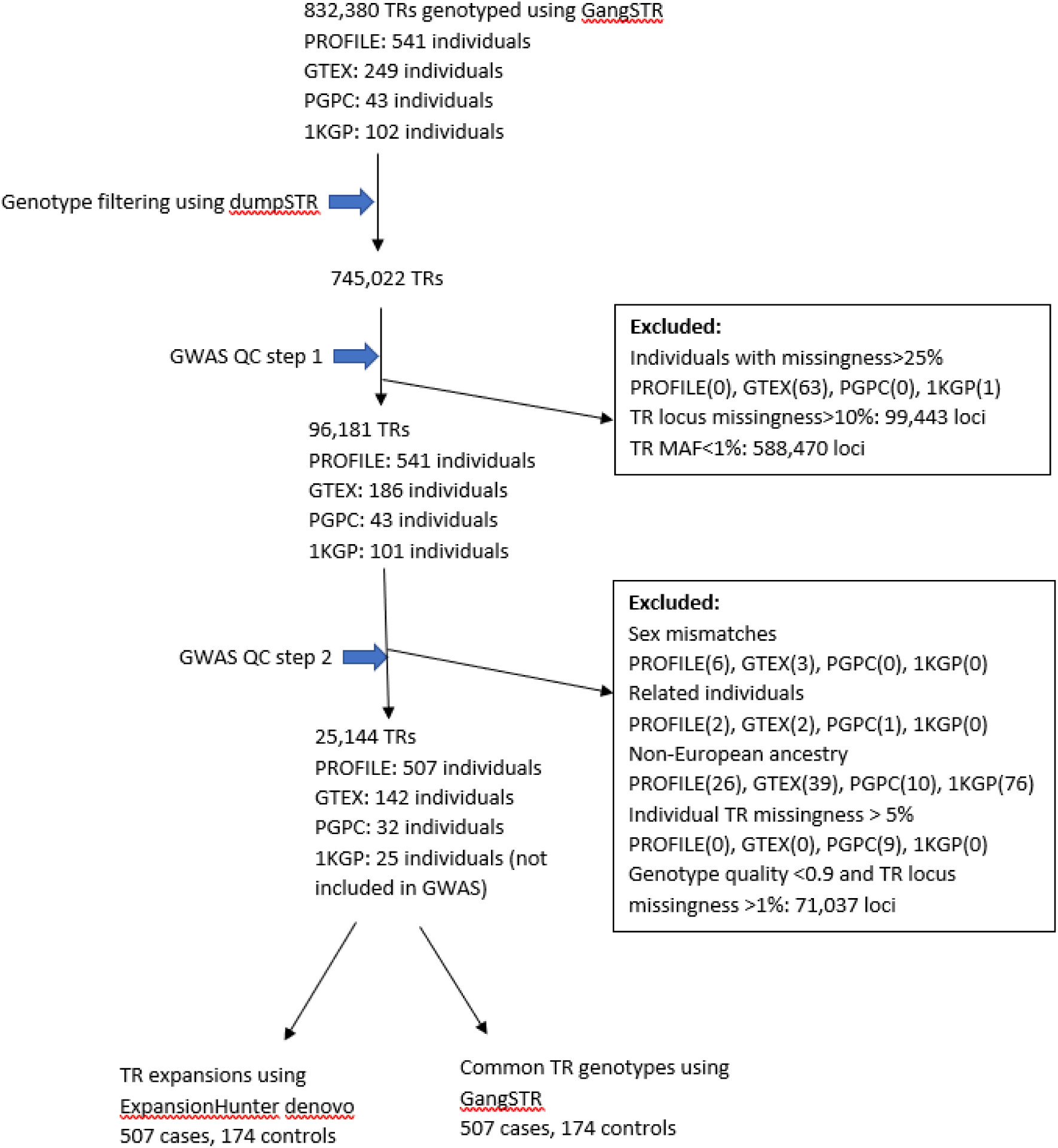
Discovery analysis.

**Supplementary Figure 2 –.**
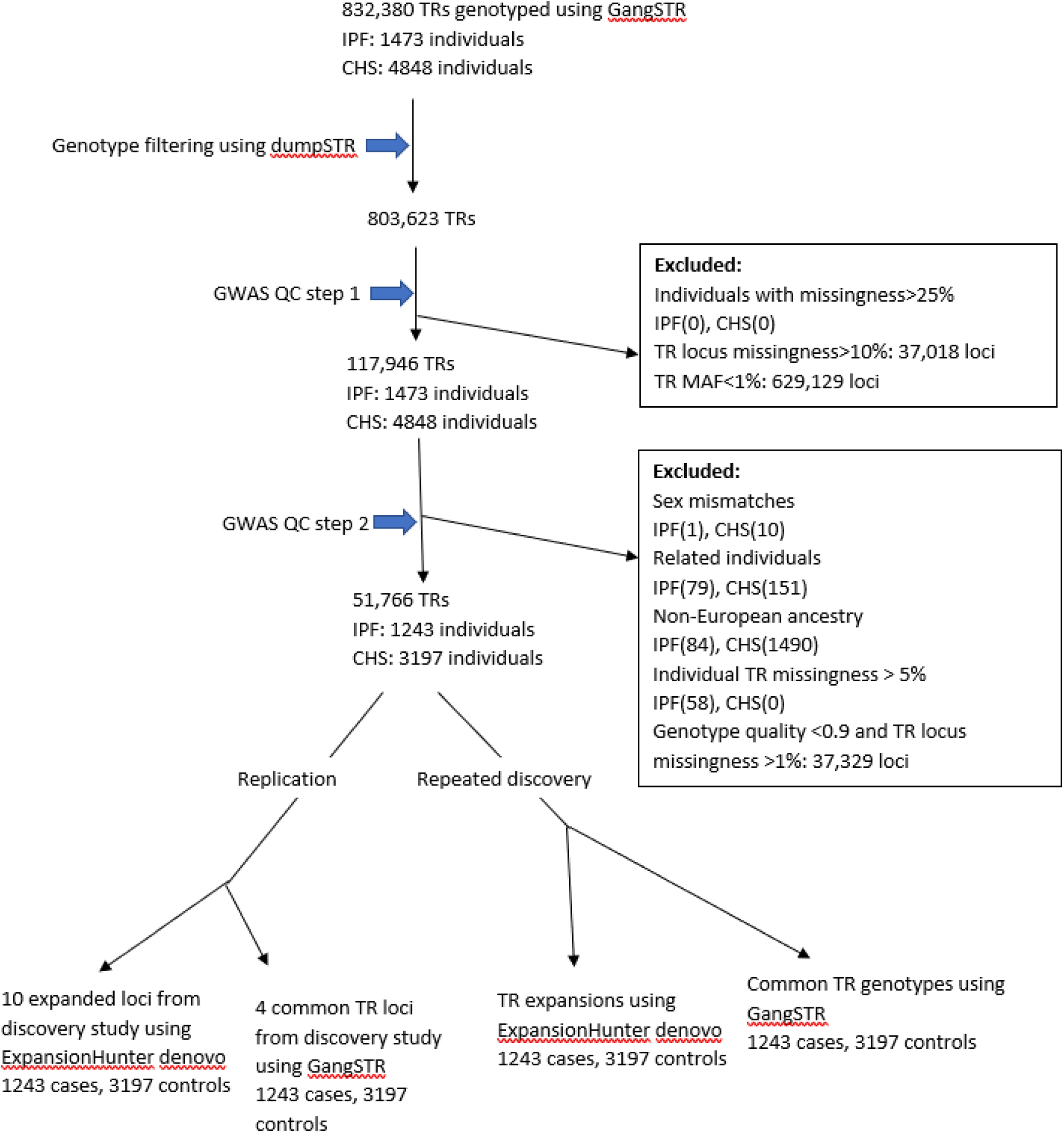
TopMed dataset replication and rediscovery analysis.

**Supplementary Figure 3.**
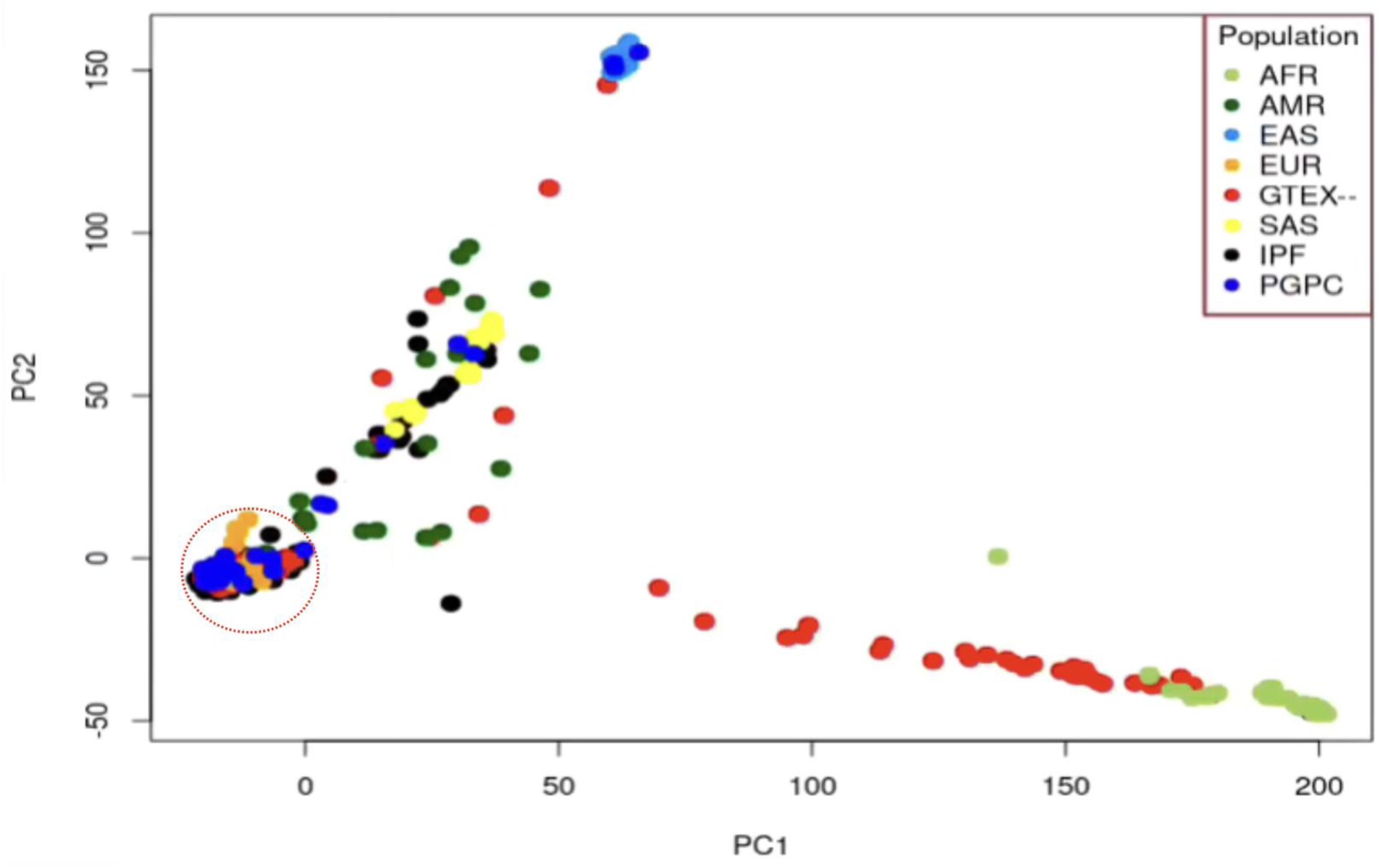
Multi-ancestry population structure PCA using STR genotype data. TR principal components of the PROFILE dataset (IPF – black), GTEX (GTEX—red), Personal Genome Project Canada dataset (PGPC – dark blue) plus 102 individuals obtained from the 1000 Genomes Project representing five major genetic ancestries i.e., African (AFR - green), European (EUR- orange), East Asian (EAS- pale blue), South Asian (SAS- yellow) and Ad Mixed American (AMR - dark-green). The population structure was determined based on clustering of sample genotypes informed by the 1KGP samples selected from a diverse population representing five major ancestries. The circle indicates individuals of European ancestry based on clustering patterns. Six median absolute deviations away from the medians of principal components 1 and 2 was used as a cut -off.

**Supplementary Figure 4.**
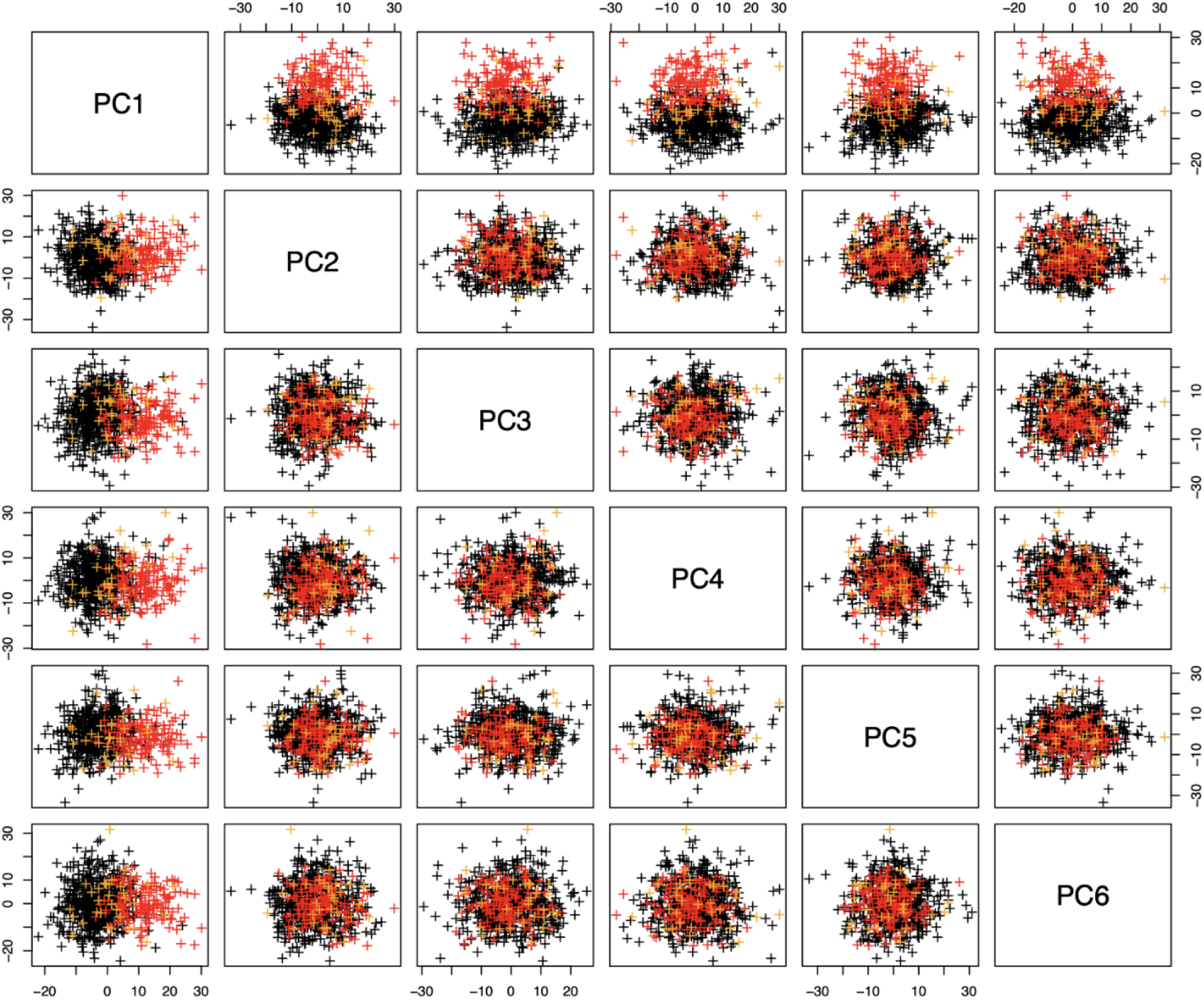
Population structure PCA using STR genotype data for discovery case-control datasets of European ancestries. Stratification across the first six principal components of 96,181 TR loci (accounting for LD by removing unique loci from 1,469 loci-pairs in LD) . The PROFILE dataset (IPF - black; n=507), GTEX PCR free (GTEX - red; n=142) and Personal Genome Project Canada dataset (PGPC - orange; n=32).

**Supplementary Figure 5.**
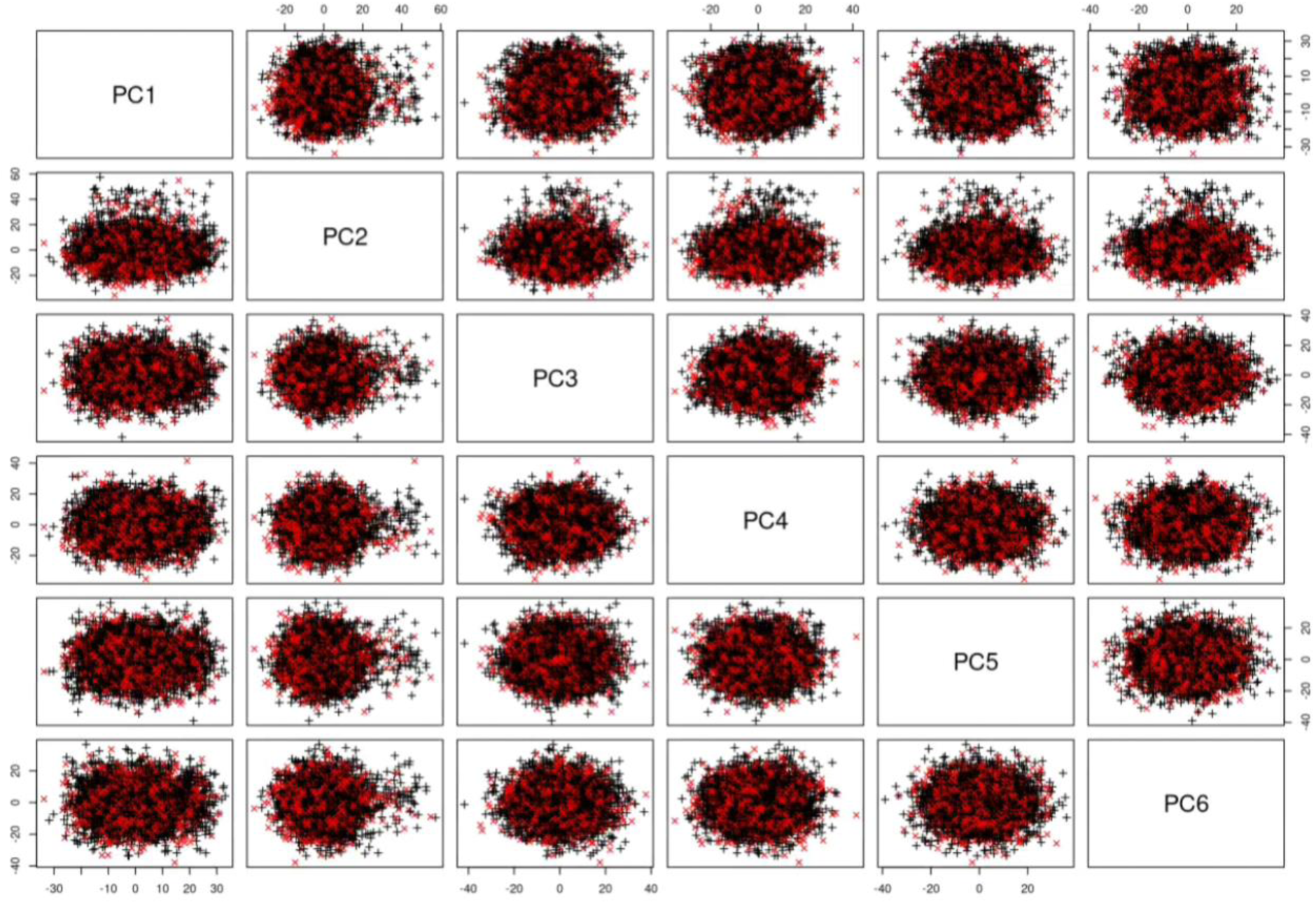
Population structure PCA for samples of European ancestry for TOPMed case-control dataset. The population structure of CHS and IPF individuals of selected European ancestry. Stratification across the first six principal components of 51,766 TR loci (accounting for LD by removing unique loci from 663 loci-pairs in LD). IPF dataset (n = 1,243) coloured in red and CHS (n=3,197) in black.

**Supplementary Figure 6 –.**
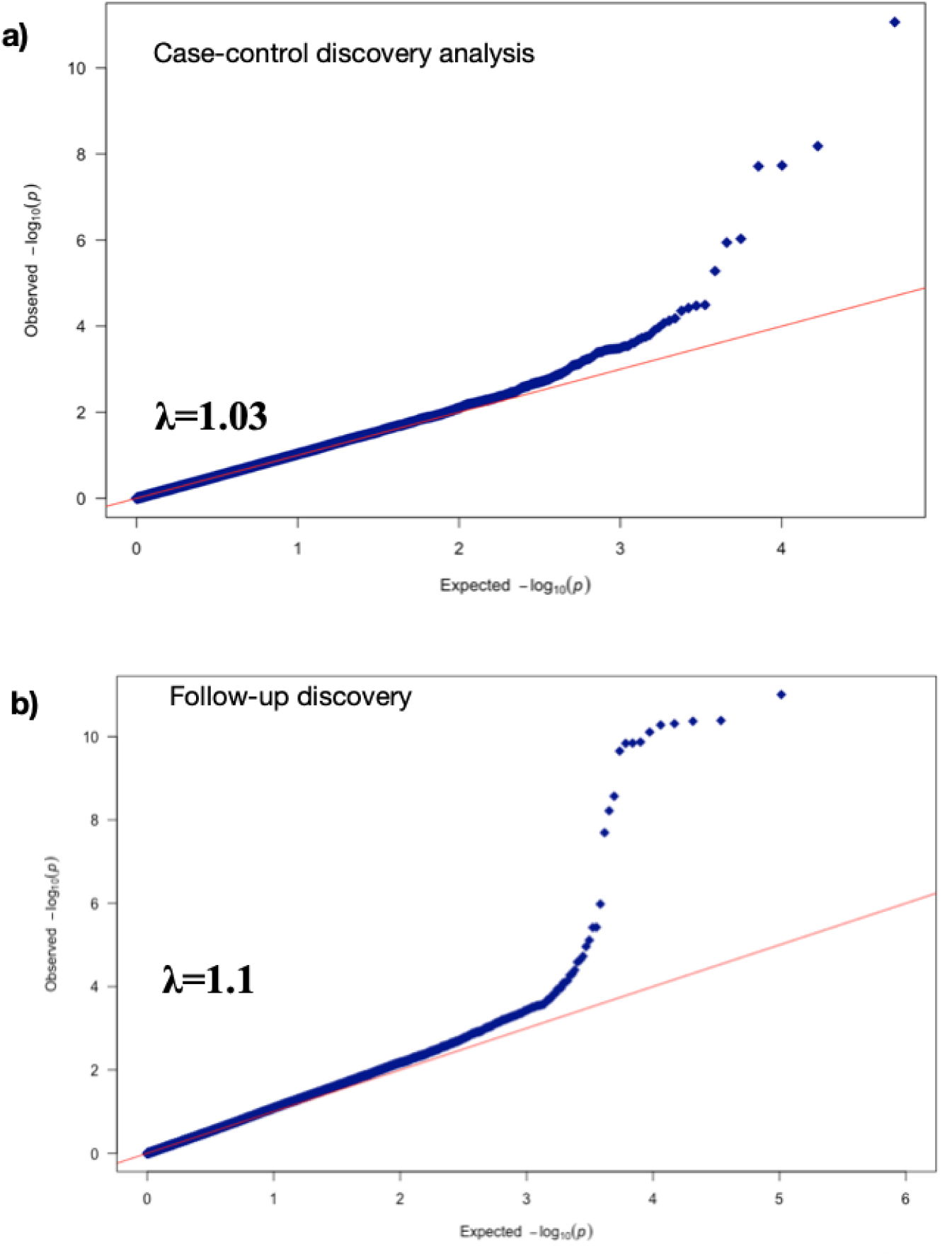
Quantile-quantile plot of expected and observed p values. a) Discovery dataset b) TopMed rediscovery dataset

**Supplementary Table 1 –.**
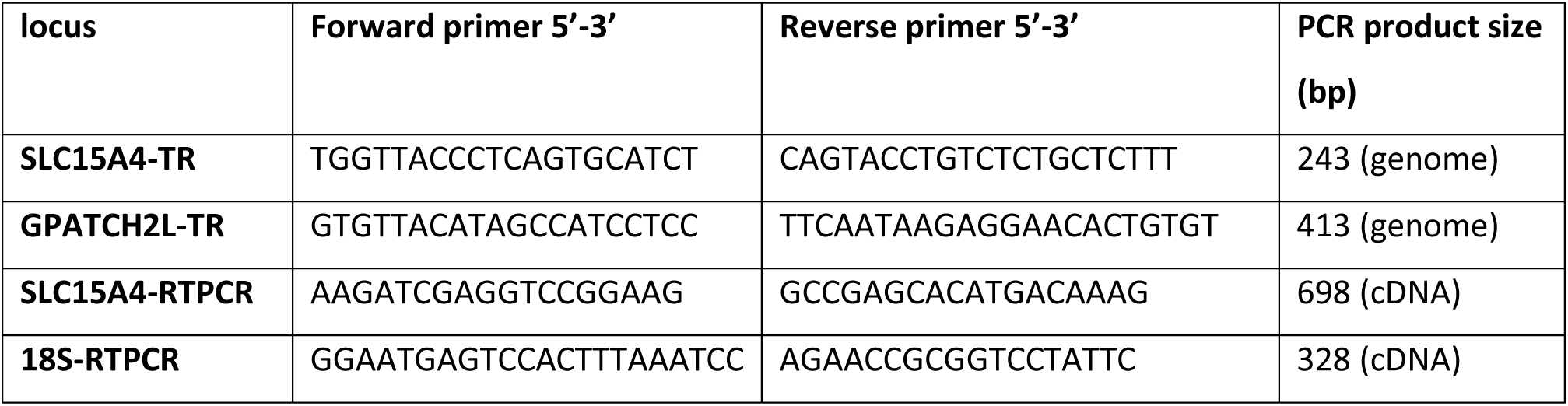
PCR primers used in this study.

**Supplementary Table 2 –.**
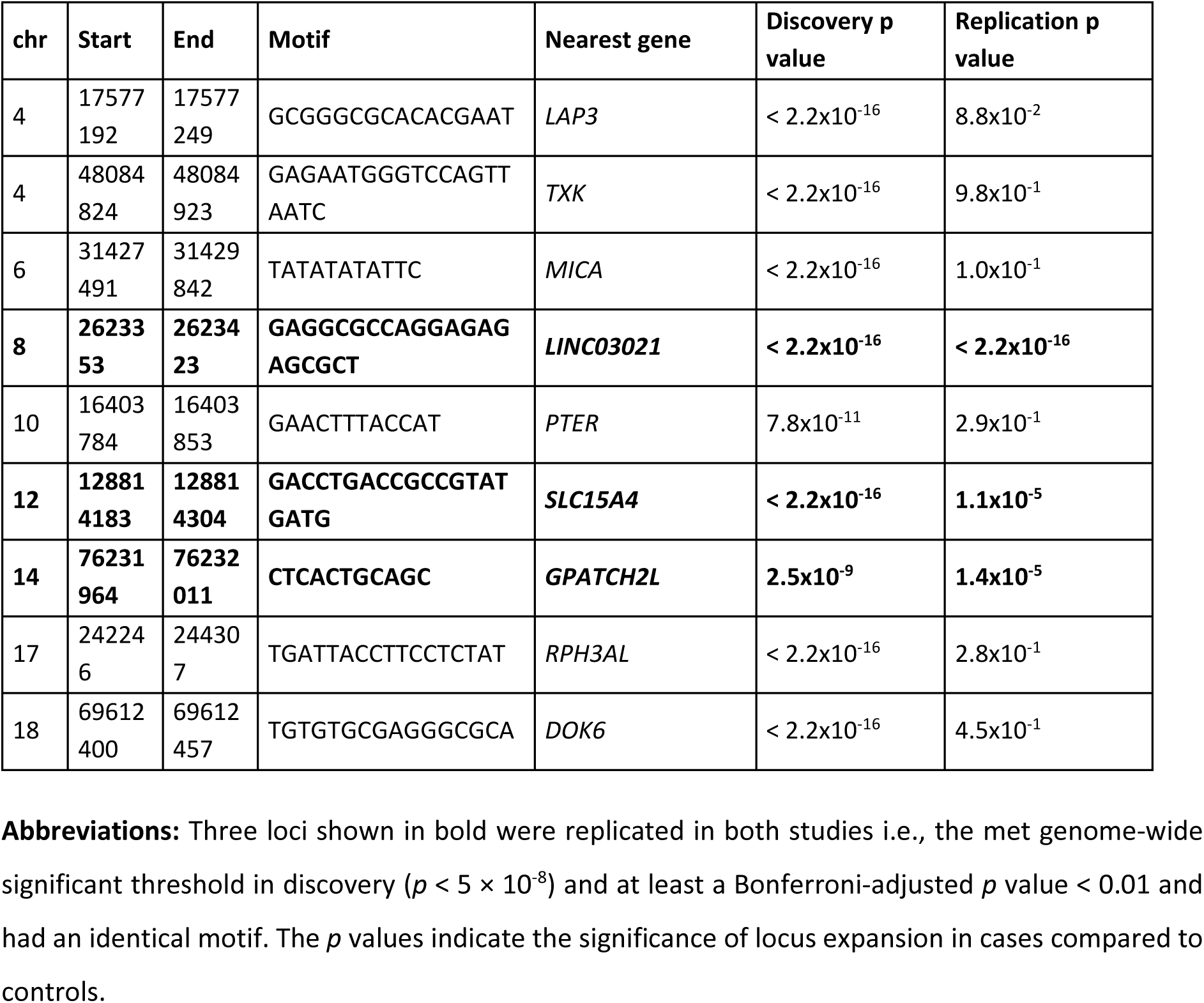
Analysis of the 10 expanded TR loci associated with IPF susceptibility.

## Notes

### Competing Interest Statement

The authors have declared no competing interest.

### Clinical Trial

NCT01134822 and NCT01110694

### Author Declarations

Ethical approval for the PROFILE study was granted by the Royal Free Hospital Research Ethics Committee (London, UK; ethics reference number 10/H0720/12) and University of Northampton Research Ethics Committee (Northampton, UK; ethics reference number 10/H0402/2) TopMed (CHS) All CHS participants provided informed consent, and the study was approved by the Institutional Review Board [or ethics review committee] of University Washington. IPF samples from the LTRC Lung Tissue Research Consortium. All LTRC participants provided written informed consent, and the study was approved by the Institutional Review Boards of the participating clinical centers (University of Michigan, Temple University, University of Pittsburgh, Mayo Clinic Rochester). Access to TOPMed data was approved by the data access committee (dbGAP study 34964)

